# Rise and Regional Disparities in Buprenorphine Utilization in the United States

**DOI:** 10.1101/19006163

**Authors:** Amir Azar R. Pashmineh, Alexandra Cruz-Mullane, Jaclyn C. Podd, Warren S. Lam, Suhail H. Kaleem, Laura B. Lockard, Mark R. Mandel, Daniel Y. Chung, Corey S. Davis, Stephanie D. Nichols, Kenneth L. McCall, Brian J. Piper

## Abstract

**Aims:** Buprenorphine is an opioid partial-agonist used to treat Opioid Use Disorders (OUD). While several state and federal policy changes have attempted to increase buprenorphine availability, access remains well below optimal levels. This study characterized how buprenorphine utilization in the United States has changed over time and whether there are regional disparities in distribution.

**Measurements:** Buprenorphine weights distributed from 2007 to 2017 were obtained from the Drug Enforcement Administration. Data was expressed as the percent change and as the mg per person in each state. Separately, the formulations for prescriptions covered by Medicaid (2008 to 2018) were examined.

**Findings:** Buprenorphine distributed to pharmacies increased about seven-fold (476.8 to 3,179.9 kg) while the quantities distributed to hospitals grew five-fold (18.6 to 97.6 kg) nationally from 2007 to 2017. Buprenorphine distribution per person was almost 20-fold higher in Vermont (40.4 mg/person) relative to South Dakota (2.1 mg/person). There was a strong association between the number of waivered physicians per 100K population and distribution per state (*r*(49) = +0.76, *p* < .0005). The buprenorphine/naloxone sublingual film (Suboxone) was the predominant formulation (92.6% of 0.31 million Medicaid prescriptions) in 2008 but this accounted for less than three-fifths (57.3% of 6.56 million prescriptions) in 2018.

**Conclusions:** Although buprenorphine availability has substantially increased over the last decade, distribution was very non-homogenous across the US.

---

Opioid agonist therapy with the medications methadone and buprenorphine, sometimes referred to as medications for opioid use disorder, (MOUD), has been shown to reduce opioid consumption, increase psychosocial functioning, and reduce drug-related and overall mortality. Systematic reviews have demonstrated a 32-69% reduction in illicit drug use, 20-60% reduction in injection drug use, and 25-86% reduction in sharing injection equipment among individuals receiving MOUD.^1,2^ Because federal law limits availability of methadone by to specialized clinics when used for the treatment of OUD, this legal status has remained stable for decades, and previous research has demonstrated that methadone distribution has remained relatively stable^3,4^ or decreased over the past decade, this paper will focus on buprenorphine.

Buprenorphine is an analogue of thebaine and functions as a mu and nociceptin receptor partial-agonist, a kappa receptor antagonist, and a delta receptor antagonist.^5^ The major metabolites are also biologically active at the four opioid receptors.^6^ Buprenorphine is eight-fold more potent as an analgesic than oxycodone and ten-fold more potent than hydrocodone.^3^ This Schedule III substance comes in several formulations: a parenteral formulation for moderate to severe pain, a transdermal patch for continuous pain management, a buccal film for moderate to severe chronic pain, a buccal film with naloxone for OUD, a subcutaneous implant designed for medication release over six-months for OUD, a long acting injection that is administered every month for OUD, a sublingual tablet (with or without naloxone) for OUD, and a sublingual film.^7^ Unlike most prescription opioids whose US use has been steadily decreasing, buprenorphine distribution increased by 75% from 2011 to 2016.^3^

There have been some concerns expressed about the safety, misuse potential, and cost of buprenorphine. Severe buprenorphine exposures including those resulting in life threatening symptoms or death reported to US poison centers increased 67% from 2011 to 2016.^8^ The US street value per mg of buprenorphine as reported to the crowdsourcing site streetrx.com was over twice that of methadone, oxycodone, and hydrocodone and four-fold higher than morphine.^9^ It is likely that the rationale for use of buprenorphine/naloxone products by persons who they were not prescribed, including occasionally by injection,^10^ is primarily to prevent withdrawal and that pleasurable effects are less common,^5,11^ particularly with repeat use. Although some prosecutors no longer accept buprenorphine possession cases because they understand that individuals using buprenorphine not prescribed are at lower risk than individuals using other opioids, there were eight-hundred arrests reported to the Maine Diversion Program from 2014 to 2017 involving buprenorphine^.12^ Buprenorphine arrests in 2017 exceeded those involving methadone, oxycodone, hydrocodone, tramadol, or morphine, combined.^13^ The buprenorphine and naloxone transmucosal film was ranked 120th in the U.S. in 2007 in sales ($70.6 million/quarter) but moved upto 28th with $415 million by the first-quarter of 2013.^14^ Racial and ethnic disparities in OUD treatment have been an persistent problem.^15,16^

There are also pronounced regional disparities in the use of prescription opioids.^13,14,17,18^ A five-fold difference was identified between the highest (Rhode Island) and lowest (South Dakoda) states.^4^ Drug overdoses are much higher in the Eastern US, compared to the Western US, although several Western states have recently seen significant increases in overdose deaths^18,19^ Half of counties, including over two-thirds in rural areas, lacked a single OUD medication provider.^20^ The prevalence of waivered physicians in 2013 differed sixteen-fold between the highest (Vermont) and lowest (Iowa) states.^21^ The objectives of this report were to extend upon existing knowledge regarding access to buprenorphine^3,4,17,20,21^ and to describe the temporal and regional changes in US buprenorphine utilization over the last decade.

## Materials and methods

### Procedures

Buprenorphine weights from 2007 to 2017 were obtained from the Automated Reports and Consolidated Ordering System (ARCOS) Retail Drug Summary Reports which are generated by the Drug Enforcement Administration’s (DEA). The year 2017 was selected as the most recent available. We utilized data for four distribution sources: pharmacies, hospitals, practitioners, and Narcotic Treatment Programs (NTPs) (values were negligible for teaching programs and mid-level providers). Population was obtained from the US Census Bureau. Examination of 2016 and 2017 was completed at a three-digit zip code level. This data source has been used in prior pharmacoepidemiological reports.^3,4,13,17^ The number of buprenorphine waivered physicians is maintained by the DEA and was obtained.^21^ Formulation information in 2008, 2013, and 2018 was procured^22^ from Medicaid. The year 2018 was selected as the most recent full-year available. The percent of the total number of prescriptions and total cost (Medicaid and non-Medicaid reimbursed) attributed to each formulation was calculated. This study was deemed exempt by the Institutional Review Board of the University of New England.

### Data Analysis

Drug weights were standardized for total state and territory population. The United States were grouped into nine divisions (e.g New England, Mid Atlantic) as defined by the Census (Supplementary Figure 1) for a regional analysis completed with Systat, version 13.1. A Pearson correlation was conducted on the association between the number of buprenorphine waivered physicians per 100K residents per state, as reported previously^21^, and per capita buprenorphine distribution. Figures were completed with GraphPad Prism, version 8.1.0. Geospatial representation was completed with QGIS, version 2.18.

**Figure 1.**
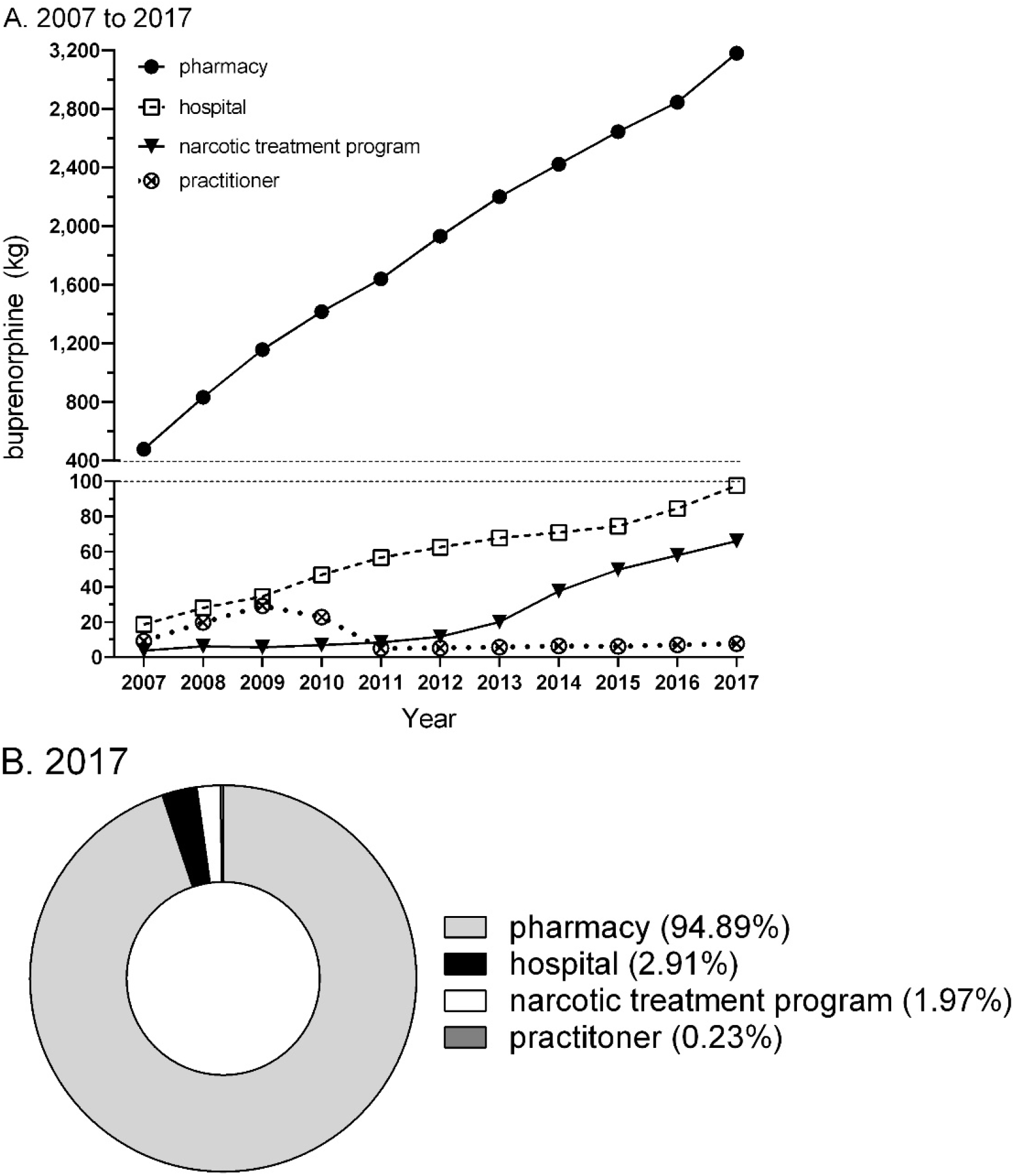
Buprenorphine (kg) from 2007 to 2017 (A) or 2017 (B) by business activity as reported to the Drug Enforcement Administration.

## Results

From 2007 to 2017, buprenorphine distributed to pharmacies increased 6.7 fold. Hospital use increased 5.2 fold (Figure 1A). The vast majority (approximately 95% throughout the study period) of buprenorphine was distributed by pharmacies (Figure 1B).

Figure 2A shows that buprenorphine utilization was not homogenous across the US. Distribution, corrected for 2017 population, differed 19.7 fold between the highest (Vermont = 40.4 mg/person) and lowest (South Dakota, 2.1 mg/person) states and was even lower (< 0.06 mg/person) in the US Virgin Islands, Guam, and American Samoa. Further analysis was completed with the states collapsed into US Census regions. Buprenorphine distribution in New England states was significantly higher than all other regions except East South Central. The Territories had significantly less than all other regions (Figure 2B). Additional examination was completed on the percent change in buprenorphine distribution from 2016 to 2017 by three-digit zip code (Figure 2C). One-third or more of the zip codes decreased (change ≤ −0.1%) distribution in Mississippi (33.3% of zip codes in the state), Colorado (35.3%), and Virginia (37.1%). Conversely, more than two-thirds of zip codes had appreciable (change ≥ +20.0%) increases in Indiana (73.7%), Arkansas (80.0%), and Connecticut (90.0%).

**Figure 2.**
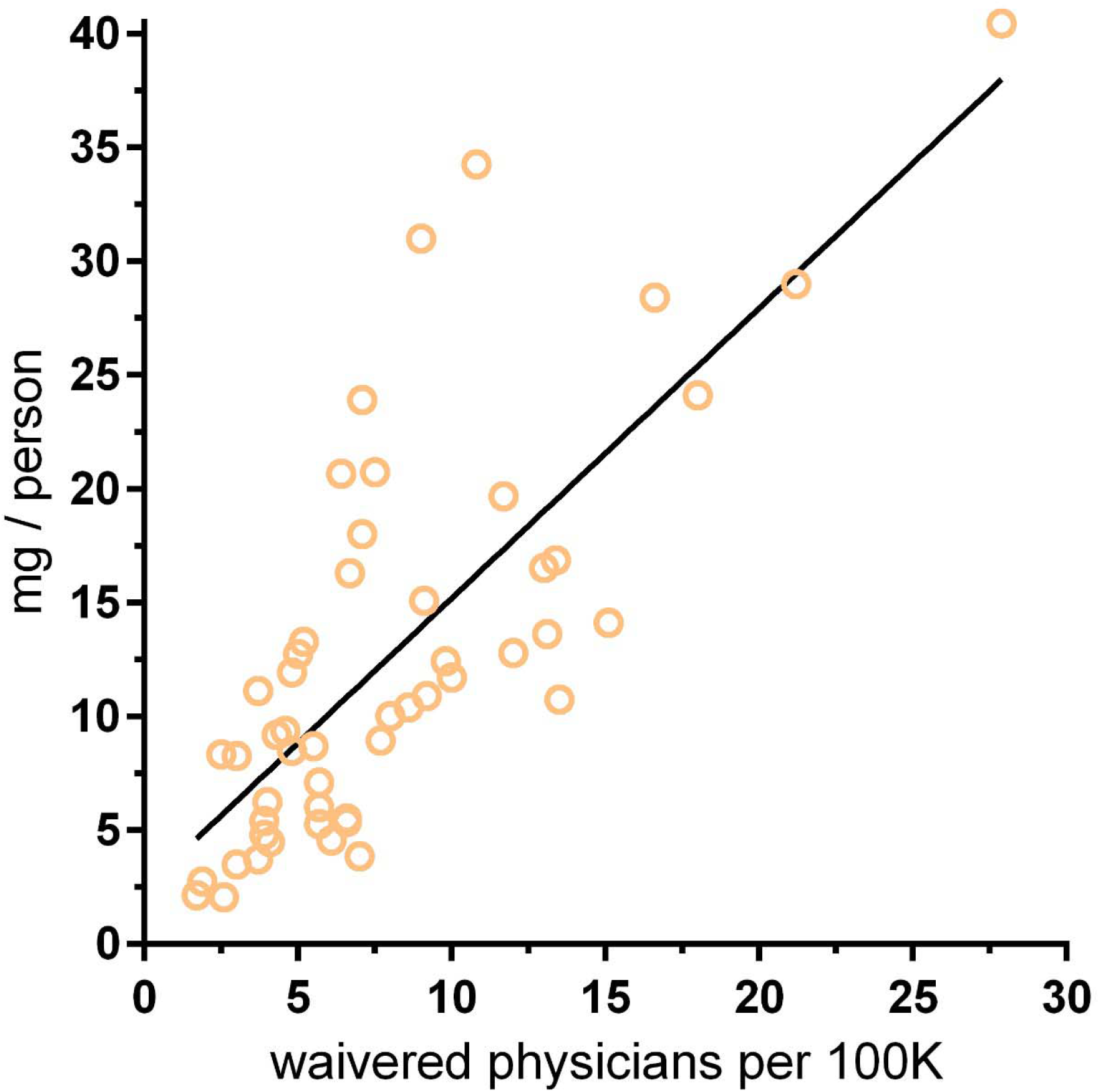
Scatterplot depicting the association (R^2^ = 0.569, *p* < .0005) between the number of buprenorphine waivered physicians per 100K population per state (from the Controlled Substances Act registrants database and reported in Table 1^21^) and buprenorphine distribution per person per state as reported to the Drug Enforcement Administration’s Automated Reports and Consolidated Ordering System.

There was a strong association between the prevalence of buprenorphine waivered physicians and buprenorphine distribution in each state (*r*(49) = 0.755, *p* < .0005, Figure 3).

**Figure 3.**
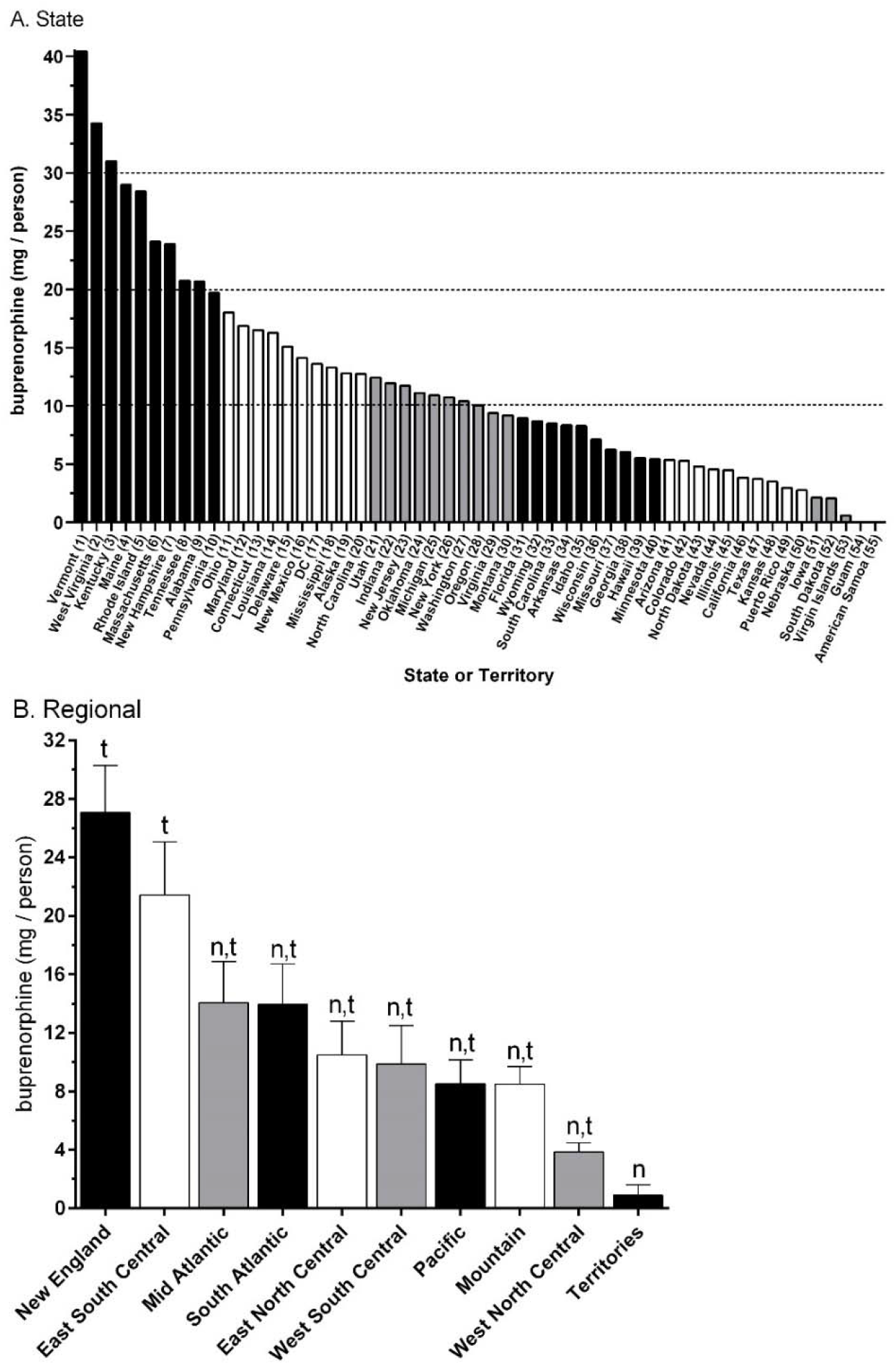
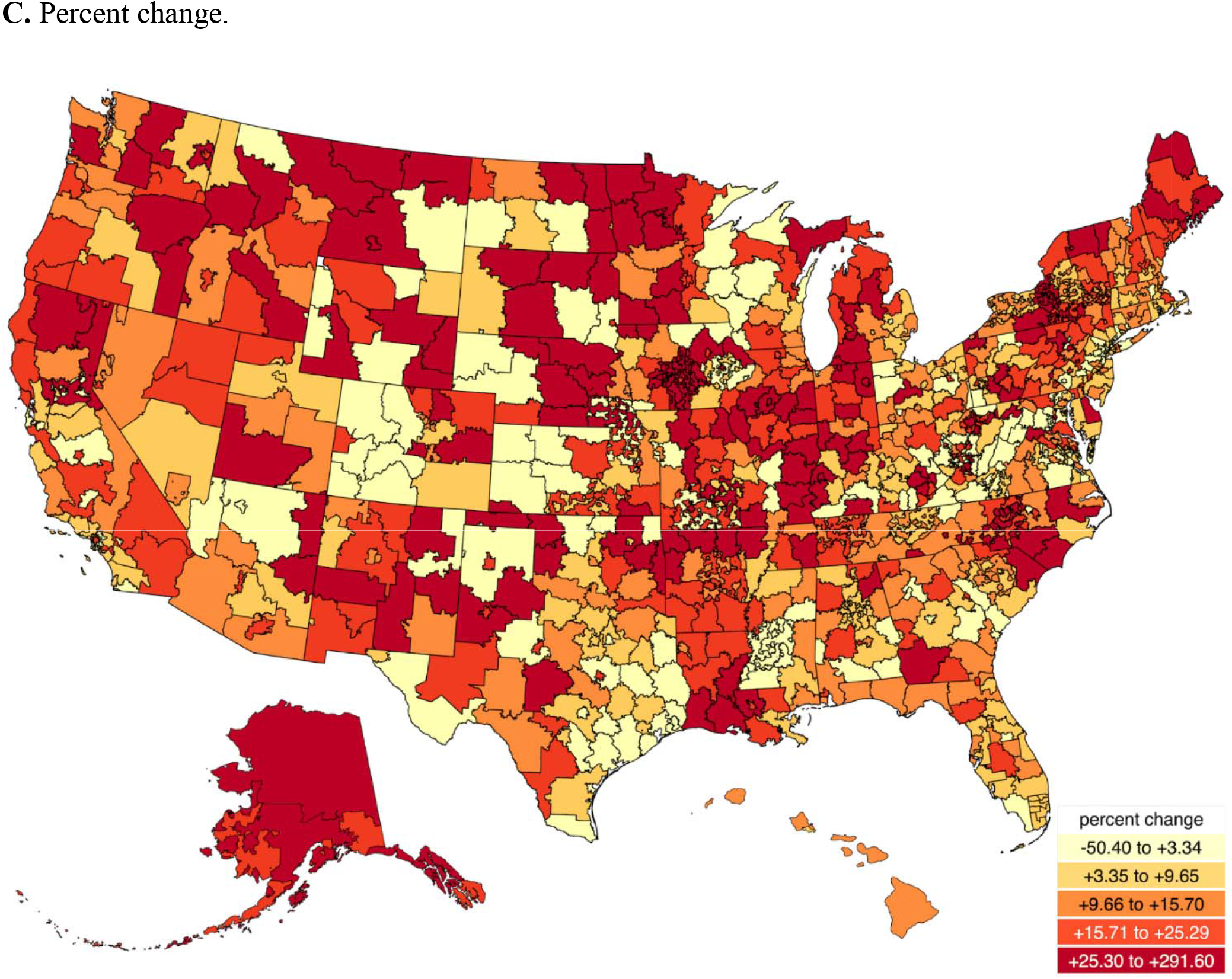
Buprenorphine use (mg / person) reported to the Drug Enforcement Administration by state (A) or US Census division (B). ^n^*p* < .05 versus New England; ^t^*p* < .05 versus the U.S. Territories (American Samoa, Guam, Puerto Rico, Virgin Islands). Percent change from 2016 to 2017 by 3-digit zip code (C).

Medicaid buprenorphine prescriptions increased five-fold from 2008 until 2013 and four-fold from 2013 to 2018. The buprenorphine and naloxone film accounted for the vast majority (92.6%) of buprenorphine formulations in 2008 and 2013 but less than three-fifths (57.3%) in 2018. Suboxone accounted for almost three-quarters of the total buprenorphine costs (1.08 billion $USD) in 2018 (Supplemental Figure 2).

## Discussion

There are two key findings of this pharmacepidemiological report. First, US buprenorphine distribution in the US has continually increased over the past decade. Second, utilization is not homogenously distributed and is highest in the eastern US. Before the year 2000, maintenance treatment of OUD involved either daily oral opioid antagonist therapy via naltrexone or opioid agonist therapy via an intensive treatment program at a methadone, or levoacetylalphamethadol (LAAM), clinic. After the year 2000, a federal law was passed that allowed office-based buprenorphine prescribing for OUD by providers who meet training and credentialing requirements including an X-waiver allowing for an additional DEA number beginning with an “X”.^23^ Buprenorphine and buprenorphine/naloxone were FDA approved for office-based treatment of OUD in 2002. However, the law permitting buprenorphine to be prescribed for OUD only permitted physicians who had received special training in addiction medicine to prescribe the medication, and severely limited the number of simultaneous patients each could treat. In 2016, the Comprehensive Addiction and Recovery Act permitted specially trained physician assistants and nurse practitioners to prescribe buprenorphine for OUD for a five-year period, and increased the patient limit. In 2018, the SUPPORT Act made permanent the provision permitting nurse practitioners and physician assistants to prescribe buprenorphine for OUD, temporarily permits certain other trained nurses to prescribe the medication, and further liberalized the patient cap.^25^

Only a limited subset of eligible physicians, nurse practitioners, and physician assistants have obtained a DEA waiver to prescribe buprenorphine for an OUD, and levels were particularly low in rural areas.^25^ Rural states like Kansas, Nebraska, Iowa, and South Dakota had the lowest buprenorphine distribution. These states also have lower rates of opioid overdose although overdose information should be interpreted cautiously because states differ in reporting procedures.^19^ Among states with very good or excellent reporting, there was a fifteen-fold difference in overdose rates between the highest (WV = 49.6 / 100K) and lowest (HI = 3.4) states.^19^ Changes in law that remove some barriers to buprenorphine access have recently been enacted, but additional changes are likely necessary to ensure that the medication is available to all those who need it.^26,27^

Four of the five US states with the most buprenorphine waivered physicians in 2013 (Vermont, Maine, Massachusetts, and Rhode Island) are in the northeast.^16^ Drug overdoses were also not uniformly distributed across the US and were more likely in New England and Appalachian areas.^28^ Eight of the ten states with the highest use of buprenorphine (Figure 2A) also had significantly elevated drug overdoses relative to the national average in 2017. ^28^ Vermont has greatly increased its capacity to OUD in the last decade with its “hub-and-spoke” model which includes observed dosing and a masters level behavioral health provider.^29^ A novel way to provide case-based education to providers about substance use disorders is through Project extension for community healthcare outcomes (ECHO) communities.^30^ Many other states in Northern New England are also working to increase capacity through Project ECHO communities. For example, New Hampshire has engaged in Project ECHO training programs to increase buprenorphine capacity among nurse practitioners (NP) and NP students through the NH Citizen’s Initiative and Maine^31^ leads many Project ECHO communities through Quality Counts.. The extremely limited use of buprenorphine in the US Territories is consistent with a prior report.^17^ The five-fold increase over the past decade in hospital buprenorphine use may reflect greater use within the emergency-room after overdoses as part of a broader effort to improve the transition to outpatient treatment for opioid addiction.^32^

Under the Affordable Care Act, all Medicaid Alternative Benefit Plans, including those offered to members of the expansion category, must provide coverage for mental health and Substance Use Disorder treatment, including buprenorphine, on parity with other medical and surgical services in compliance with the Mental Health Parity and Addiction Equity Act. Recent evidence demonstrates that compared to non-expansion states, Medicaid expansion states experienced increases in overall prescriptions for, Medicaid-covered prescriptions for, and Medicaid spending on evidence-based OUD pharmacotherapies, particularly buprenorphine and naltrexone.^33-36^

Some limitations and future directions are noteworthy. First, although buprenorphine distribution was increasing overall, examination of individual 3-digit zip codes indicates that the opposite pattern may be occurring in some rural areas. Other data sources will be needed to identify and monitor specific populations^15,16^ (e.g. criminal justice) where there continue to be access barriers. Second, the DEA data source, although comprehensive, does not allow for discrimination between different buprenorphine formations or their transportation across state lines (i.e. mail order pharmacies). Medicaid complements ARCOS in that it provides detailed information on formulations but only covers one-fifth of the US population. These data sources track distribution to pharmacies and hospitals but do not differentiate subsequent medical^3,4,17^ versus nonmedical use.^5,9,10,12,13^

In conclusion, although this report identified pronounced increases in buprenorphine distribution, this study also discovered substantial regional differences. Further research is warranted on strategies to optimize OUD pharmacotherapy access and long-term treatment outcomes.

## Data Availability

Original data is available online.

https://www.deadiversion.usdoj.gov/arcos/retail_drug_summary/index.html

https://www.medicaid.gov/medicaid/prescription-drugs/state-drug-utilization-data/index.html

## Acknowledgements

Software to complete this project was provided by Husson University School of Pharmacy and the NIEHS (T32 ES007060-31A1). BJP was a Fahs-Beck Fellow and supported by the Center of Excellence, Health Resources Services Administration (D34HP31025). An earlier version of this study was presented at the International Society for Pharmacoepidemiology’s 35^th^ annual meeting.

